# Rate and predictors of Treatment Failure among pediatric population taking Highly Active Antiretroviral Therapy in Ethiopia

**DOI:** 10.1101/19005538

**Authors:** Yimam Getaneh, Ajanaw Yizengaw, Agajie Likie, Mulusew Getahun, Altaye Feleke, Eleni Kidane, Amelework Yilma, Achamyeleh Mulugeta, Tezera Moshago, Yibeltal Assefa

## Abstract

**Background:** Though the unprecedented global effort at scaling up universal access to antiretroviral therapy (ART) has decreased the progression of HIV, treatment failure (TF) among pediatric patients receiving ART against human immunodeficiency virus (HIV) is becoming a global public health concern which may impact on treatment outcome. Thus, the aim of this study was to determine the rate and predictors of treatment failure (TF) among HIV-infected pediatric patients taking ART in Ethiopia.

**Methods:** A prospective and retrospective follow-up study was conducted from March 2016 to 2017. Retrospective clinical and laboratory data were captured from patients’ medical record. Socio-demographics and explanatory variables of participants were collected using pre-tested structured questionnaire and study participants were followed for three to six month after baseline viral load has been done to classify virologic failure (VF). TF was ascertained from population who virally failed with the denominator of population taking ART. Chi-square test and multiple logistic regressions were conducted to assess predictors TF. Statistical significance was set at P-value less than 0.05.

**Results:** A total of 554 pediatrics patients taking ART from 40 selected health facilities were included in the study. Viral load suppression (VLS) (VL<1000 copies/ml) among pediatric population taking ART in Ethiopia were found to be 344 (62.1%). From those who was not virally suppressed at baseline of the study 210 (37.9%), 99 (51.6%) were re-suppressed after three to six month of enhanced adherence and counseling, leading the overall virologic failure (VF) among pediatric population taking ART in Ethiopia to be 93 (17.3%). The mean CD4 count was improved from 490 cells/ml at ART initiation to 921 cells/ml after 80 months of ART exposure. Moreover, the clinical outcome was improved from 42% to 89% at ART initiation and after 80 month of ART experience. CD4 count, clinical stage, Hemoglobin and weight were found to be predictors of VF. Moreover; family HIV and disclosure status, duration on ART, age, being orphan, stigma and medication adherence have significant association with VF.

**Conclusions:** The low level of VLS (62.1%) and the high level of VF (18.3%) could explain the challenge on the national ART program among pediatric population. The significant improvement on immunologic and clinical outcome could indicate the success of ART on treatment outcome among pediatric population. CD4 count, clinical stage, Hemoglobin and weight could be good predictors of TF among pediatric population. Improving disclosure status, stigma and medication adherence could improve the treatment outcome of pediatric population taking ART in Ethiopia.

## Background

The global scaling up of treatment and care for people living with human immunodeficiency virus (PLHIV) has led to a 43% decline in new HIV pediatric infections since 2003, with 330,000 newly infected children in 2011[1]. Despite efforts to expand access to antiretroviral therapy (ART), only 28% of eligible children have received it [2]. Expansion of early HIV diagnosis coverage, prompt ART initiation and better retention in care remain major goals [3], [4], and the lack of laboratory monitoring frequently observed in low and middle-income countries (LMIC) should not represent a barrier to ART distribution in children [2]–[6]. In Ethiopia, the total number of pediatric population living with HIV in 2018 was estimated to be 56,514 of which 2,994 were new infections[7]. Very few pediatric patients among these are started on ART regimens in Ethiopia.

Although virological failure is widely considered the criterion standard to detect treatment failure, clinical and immunologic parameters are often the only criteria available in LMIC. CD4 cell monitoring has been shown to be a poor predictor of virological failure in treatment experienced children, particularly when severely immune-compromised [8]–[13]. Studies in LMIC have reported high rates of virological suppression in children up to 5-6 years after treatment initiation; however, treatment failure rates of 10-34% were observed among children after 2-3 years of ART [14]–[19].

Previous studies have also evaluated the patterns associated with switching from first line ART regimes to second line ART regimes; however, studies that evaluate the rate and predictors treatment failure in Ethiopia and Africa at large are scarce. Hence, this study reported rate of first line ART treatment failure and predictors which will help to guide program experts and decision makers working on the ART program and clinicians to evaluate patients on ART in every visit for treatment failure and contributing factors. The study result can also be used as a baseline data for subsequent studies.

## Methods

### Study design and population

A retrospective and prospective follow-up study was conducted across the nationally representative health facilities (HFs) in Ethiopia. Baseline VL testing was done followed by second round VL testing after three to six months of intensive adherence counseling for patients with VL >1000 copies/ml at baseline to determine treatment failure (TF) as defined by WHO-2013 [20]. Pre-established tools and questioner were also used to assess determinants of TF. According to the WHO recommendation, selection of 40 HFs is sufficient for nationally representative virologic failure (VF) [21], [22]. Magnitude of VF among pediatric HIV-1 infected patients in Gondar university hospitals in Ethiopia is reported as 18.2% [16]. A confidence interval of half-width of ±5% has been used as appropriate compromise between feasibility and precision with 95% Confidence Interval (CI). Moreover, numbers of adults on ART within the HF across each stratum by the end of 2014 have been independently considered and calculated using the formula;

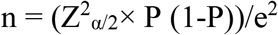

Where;

Z = value from standard normal distribution corresponding to desired confidence level (Z=1.96 for 95% CI)

P= Expected true proportion (0.182)

e= Desired precision (0.05)

Hence the assumed samples size was calculated to be 229. Considering the response rate of 85%, laboratory failure rate 7% and design effect of 2, the required sample size was calculated to be 559.

Immunologic failure (IF), VF and clinical failure (CF) were considered to be dependent variables whereas socio-demographic variables, duration on ART, variable related to patient social behavior (disclosure, use of treatment assistant, use of memory aids, use of alternative medicine, use of alcohol and substance abuse, missed appointments), knowledge and perception on HIV and ART (knowledge and information on ART, Perception of treatment) and ART service delivery environment (waiting time, distance to the clinic, quality of care, trust in health care, providers, pill burden) were independent variables.

## Data collection

Data were collected in two rounds, to classify the study population whether confirmed treatment failure or not per the WHO classification [20]. Base line data were captured for the study participants from March to August, 2016. Second-round data collection was specific for the study participants with baseline viral load >1000 copies/ml since December, 2016 to March, 2017.

### Types of data source

#### Primary data source

Pre-established questioner, used to capture data related with medication adherence, knowledge, attitude and perception on ART, service delivery environment and other demographic variables.

#### Secondary Data source

Secondary data were collected from the review of participants‘medical record. These variables included medical history at three points; by the time the participant start ART, the latest record prior to the data collection and record during the data collection. Some of the key variables were CD4^+^ T-cell count, Clinical status per WHO classification, Medication adherence history.

#### Blood sample collection

Five milliliter blood sample was collected from the study participants during their attendance of the health facility for their appointment follow up. Plasma was separated after centrifugation and transported according to the standard operating procedure for sample collection, transport and tracking.

#### CD4^+^ T-cell count

CD4^+^ T-cell count was conducted at the health facility laboratory using either BD FACS Count ^™^ or BD FACS Calibour ^™^ (Becton Dickinson, USA).

#### Viral load testing

VL testing was conducted at base-line of the study and repeated for the study participants with baseline viral load >1000 copies/ml for the second round.

Viral load testing was conducted at the regional laboratories of the country using Abbott m2000sp system (Abbott Laboratories, Abbott Park, IL, USA) and COBAS® AmpliPrep/COBAS® Taq Man® HIV-1 Test, v2.0.

## Statistical analysis

The cumulative magnitude of VF was estimated from the proportion of children with VL>1000 copies/ml both at baseline and second round of the study. Similarly, magnitude of immunologic and clinical failure were determined as per WHO definition [20].

Factors associated with VF was evaluated by comparing variables among adult who failed with those who never failed using the chi-square test for categorical data, and using student-t test for continuous variables. Logistic regression was done to determine the factors contributing to VF. The model were then built by dropping the most insignificant factor one at a time with factors whose P<0.05 were taken to be the factors that were independently associated with VF. All analyses were done using SPSS version 20.

### Operational definitions

|  |  |
| --- | --- |
| Adherence: | The degree to which the person's behavior corresponds with the agreed recommendations from a health care provider. |
| Early stage ART experience: | Population with ART experience from 9 to 15 months |
| Mid-level ART experience: | Population with ART experience from 16 to 47 months |
| Advanced ART experience: | Population with ART experience with at least 48 months |
| Clinical Failure: | New or recurrent clinical event indicating severe immunodeficiency (WHO clinical stage 4 condition) after 6 months of effective treatment |
| Immunologic Failure: | CD4 count falls to the baseline (or below) or persistent CD4 levels below 100 cells/mm<sup>3</sup> |
| Virologic failure: | Plasma viral load above 1000 copies/ ml based on two consecutive viral load measurements after 3 months, with adherence support |
| Viral suppression: | Plasma viral load above 1000 copies/ ml based on one viral load measurement |

## Result

Between March 2016 and 2017, 554 HIV-infected children less than 15 years old who were on ART in 40 selected health facilities in Ethiopia were included in this study.

### Characteristics of the study population

Demographically, 269 (48.6%) and 274 (51.4%) were female and male respectively. Moreover, 489 (88.3%) and 57 (11.7%) were originally from urban and rural respectively. More than half 325 (58.9%) of the study participants were above the age of 120 months followed by 61-120 months accounted for 198 (35.9%). More than half of the study participants 235 (57.6%) had at least 48 month of ART experience while 126 (30.9%) and 47 (11.5%) had 15-47 and less than 15 months ART experience, respectively (Table-1).

**Table-1:** Socio-demographic characteristics of HIV-infected pediatric population receiving ART in Ethiopia from March 2016 to 2017

| Variable |  | Frequency | Percent |
| --- | --- | --- | --- |
| Gender | Female | 269 | 48.6 |
|  | Male | 274 | 51.4 |
| Residency | Urban | 489 | 88.3 |
|  | Rural | 57 | 11.7 |
| Duration on Treatment (Months) | ≤ 14 | 47 | 11.5 |
|  | 15-47 | 126 | 30.9 |
|  | 48+ | 235 | 57.6 |
| Age (Months) | ≤ 60 | 29 | 5.3 |
|  | 61-120 | 198 | 35.9 |
|  | 120+ | 325 | 58.9 |

### Rate of treatment failure among pediatric population taking ART in Ethiopia

VLS among pediatric population taking ART in Ethiopia was 344 (62.1%). Study participants whose viral load >1000 copies/ml (N=210) were followed for three to six month with adherence counseling. With 18 (8.5%) dropout, 99 (51.6%) were re-suppressed and the remaining 93 (48.4%) of those who were virally not suppressed at baseline were not re-suppressed and classified as confirmed VF. Hence, the overall VF among pediatric population taking ART in Ethiopia was found to be 93 (17.34%) (Table-2). Participants at clinical stage I were significantly improved from 26% at ART initiation to 73% after average 39 months of ART experience. Clinical outcome was improved from 42% to 89% at ART initiation and after 80 month of ART experience. Moreover, the mean CD4 count was improved from 490 cells/ml at ART initiation to 921 cells/ml after 80 months of ART exposure (Figure-1).

**Figure-1:**
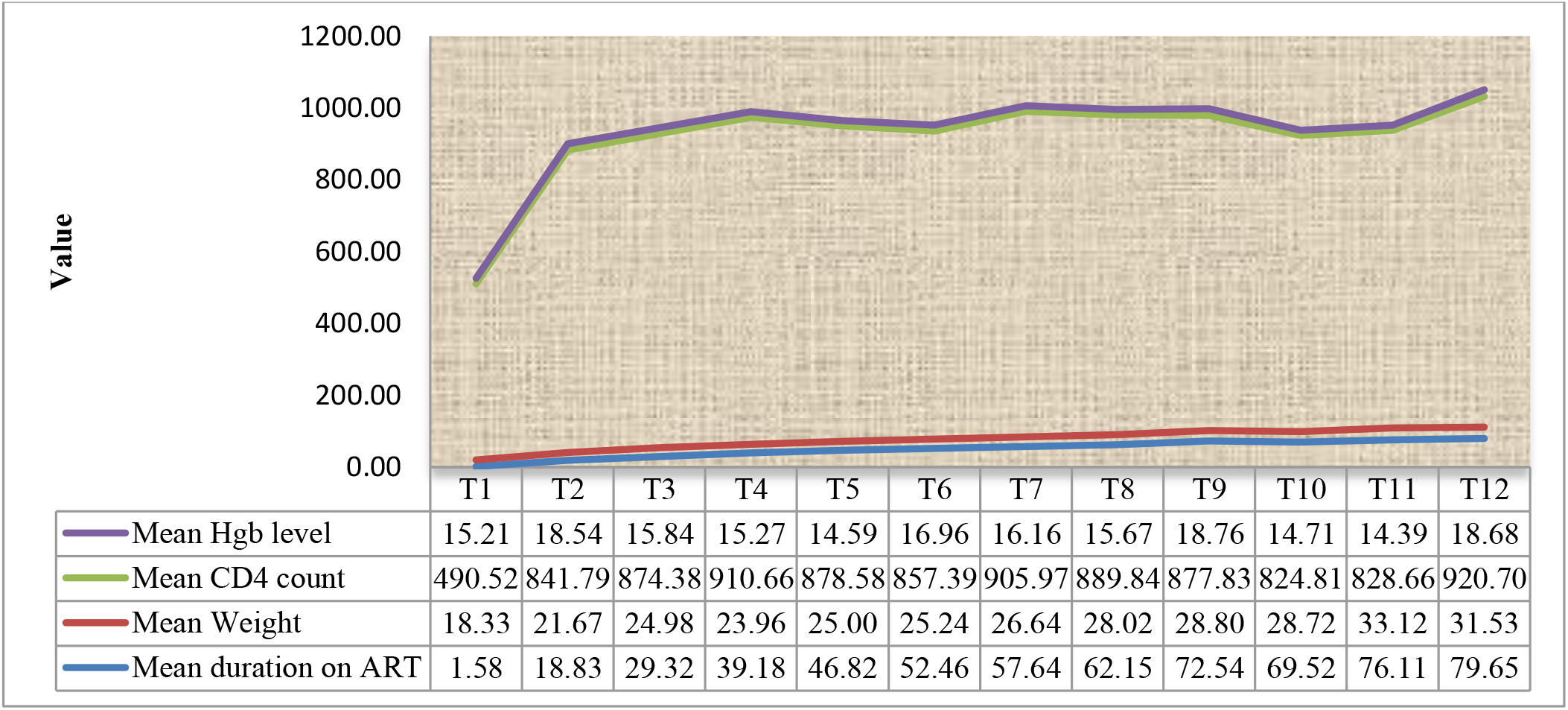
Treatment outcome among pediatric population taking ART in Ethiopia

**Table-2:** Treatment failure among pediatric population taking First line ART in Ethiopia from March 2016 to 2017

|  | Variable | Frequency | Proportion (%) |
| --- | --- | --- | --- |
| WHO Clinical stage* | I | 137 | 26.0 |
|  | II | 161 | 30.6 |
|  | III | 195 | 37.1 |
|  | IV | 33 | 6.3 |
| VLS | Not suppressed | 210 | 37.9 |
|  | Suppressed | 344 | 62.1 |
| VF | Not virally failed | 99 | 51.6 |
|  | Virally failed | 93 | 48.4 |
| Adherence (VAS)* | poor | 20 | 3.7 |
|  | Fair | 26 | 4.8 |
|  | Good | 495 | 91.5 |
| CD4 count* | ≤250.00 | 167 | 33.3 |
|  | 250.01-500.00 | 161 | 32.1 |
|  | ≥500.01 | 173 | 34.5 |
| Regimen* | 1B | 1 | 0.40 |
|  | 1C | 51 | 20.16 |
|  | 1D | 15 | 5.93 |
|  | 1E | 15 | 5.93 |
|  | 1F | 8 | 3.16 |
|  | 1G | 9 | 3.56 |
|  | 1H | 4 | 1.58 |
|  | 2H | 1 | 0.40 |
|  | 4A | 3 | 1.19 |
|  | 4B | 1 | 0.40 |
|  | 4C | 103 | 40.71 |
|  | 4D | 34 | 13.44 |
|  | 4E | 3 | 1.19 |
|  | 4F | 1 | 0.40 |
|  | 4G | 4 | 1.58 |

\*Data at ART initiation

### Predictors of pediatric treatment failure among population taking ART in Ethiopia

This study revealed that, there is significant association between VLS with maternal HIV status, being orphanage, duration on treatment, CD4 count at ART initiation and residency (P=0.046, 0.049, 0.001, 0.001 and 0.032) (Table-3). Living with mother or father was found to be preventive for viral suppression (AOR=0.51, 95% CI: 0.248-4.47). Similarly, Early stage ART experience was also found to be a risk for VLS (AOR: 1.86, 95% CI: 0.42-3.31). CD4 count and WHO clinical stage were also predictors for treatment failure (Table-4). The mean level of Hgb was improved from 15.21 at ART initiation to 18.68 after 80 months of ART experience. Moreover, the mean CD4 count and weight was improved from 490.52 and 18.33 at ART initiation to 920.70 and 31.53 after 80 months of ART ignition, respectively (Figure-1).

**Table-3:**
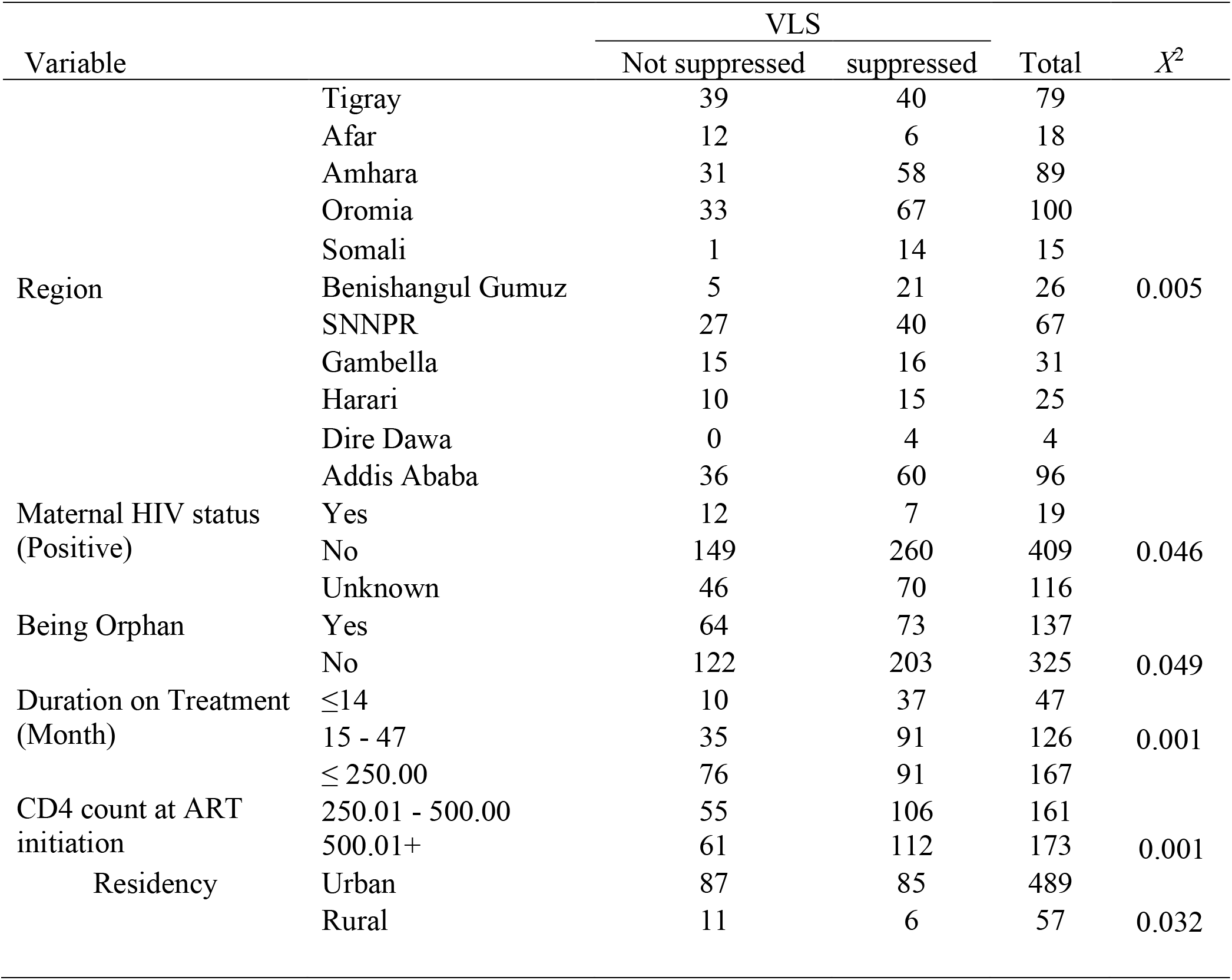
Factors associated with Pediatric treatment failure in Ethiopia

**Table-4:** Predictors of Pediatric treatment failure in Ethiopia

| Variable |  | AOR | Sig. | 95% CI |  |
| --- | --- | --- | --- | --- | --- |
|  |  |  |  | Lower | Upper |
| Family status | Living with Family (Mother or Father) | 0.51 | .03 | 0.248 | 0.77 |
|  | Living with Care Giver | 3.04 | .06 | 1.95 | 6.28 |
|  | Living with Family (Uncle, Ant...) | 2.04 | .09 | 1.37 | 4.45 |
|  | Orphan | 3.22 | .06 | 2.46 | 6.59 |
|  | Other | Reference |  |  |  |
| Mother on ART | Yes | .61 | .65 | 0.35 | 0.92 |
|  | No | Reference |  |  |  |
| Disclosure | Yes | .91 | .06 | .061 | 0.98 |
|  | No | Reference |  |  |  |
| Duration on ART (in Months) | ≤ 14 | 1.86 | .012 | 1.41 | 3.31 |
|  | 15- 47 | 1.18 | .006 | 1.13 | 2.02 |
|  | 48+ | Reference |  |  |  |
| CD4 Count (Cells/ml) | ≤ 250 | .71 | .107 | .154 | 1.58 |
|  | 250- 500 | 1.02 | .022 | 1.46 | 1.90 |
|  | 500+ | Reference |  |  |  |
| WHO clinical stage | I | 1.58 | .044 | 1.42 | 3.12 |
|  | II | .70 | .333 | .721 | 2.12 |
|  | III | .90 | .190 | .446 | 2.24 |
|  | IV | Reference |  |  |  |

## Discussion

From this study, VLS among pediatric population taking ART in Ethiopia were found to be 344 (62.1%). Of those who was not virally suppressed at baseline of the study 210 (37.9%); 99 (51.6%) were re-suppressed after three to six month of enhanced adherence and counseling, leading VF among pediatric population taking ART in Ethiopia to be 93 (17.32%). This finding is consistent with a study conducted in Tanzania which revealed viral suppression as 68.4% [10]. However, this contradicts with other study conducted at Rohde Island which showed 43% viral suppression among pediatric population[14]. This study revealed that pediatric TF is a programmatic concern since there is significant difference with VLS among pediatric versus adult in Ethiopia which was reported as 88.1%[23]. Immunologic failure at base line of this study was 185 (33.3%) at ART initiation and improved to 39 (7%) after 80 month of ART experience. This contradicted with a study conducted at University of Gondar which revealed magnitude of IF at ART initiation be 10%[12]. From retrospective cohort study done in Nigeria the rate of first line regimen failure was 18.8%[26].

In this study, Immunologic failure was found to be 33.3% at base line of ART initiation while the majority of (67.7%) were at stage clinical stage II/III at baseline of ART iniation which has been significantly improved after 80 month of ART initation. This contradicted with a retrospective cohort study done at Jimma University Specialized hospital, among children on first line ART, clinical treatment failure and immunologic treatment failure were diagnosed in 6.2% and 11.5% respectively[27]. From the retrospective cohort study conducted in Addis Ababa, there were 14.1% children with HIV/AIDS who had evidence of first line ART failure of which 5.9% had clinical treatment failure, 6.7% immunologic failure and 1.5% developed both immunologic and clinical failure [12]. From all studies above the prevalence of the ART treatment failure was not-comparable except that of Ugandan study which was higher and explained by the nature of study being case control with possible risk factors. From study conducted at Addis Ababa, out of all children with first line ART failure, only 24 (14.4%) were identified. The mean time of detection of treatment failure was 19.7 months (SD = 14 months) and the mean time to switch to second line ART regimen, for those switched, was 24 months [12].

The detection rate was higher in the current study which can be due to the high prevalence of treatment failure and the increased awareness for treatment failure as time goes. In this study duration of ART (p=0.049) was independent factor that increase the risk of ART treatment failure. Similar to the study conducted in Medical College and Research Institute of Bangalore, duration on ART for more than 3 years (P=0.0436) was associated with immunological failure[28]. In multiple regression, duration on ART, age and CD4 count (lowest ever) on treatment were predictors of immunological failure in these patients[28]. As it can be seen from these studies the chance of ART treatment failure was increasing as duration of ART increases which is consistent with this study. This study also showed that base line CD4 less than 250 cells/µl (p<0.001) were independent factors for ART treatment failure. In Medical College and Research Institute of Bangalore, low CD4 counts (<100cells/μl) at start of ART (P=0.0261), less than 50% gain in CD4 count (P=0.048) after one year of start of ART and duration on ART for more than 3 years (P=0.0436) were associated with immunological failure[28]. In multiple regression, duration on ART, age and CD4 count (lowest ever) on treatment were predictors of immunological failure in these patients [16]. From the studies conducted in Nigeria and Cambodia risk factors for ART treatment failure were ARV exposure and sever immunosuppression before start of ART [26]. From a study conducted in Kenya, the factors were base line CD4 below 50 cells [29]. From these studies, the risk factors including low base line CD4 and poor adherence were found to be significant associated factors for ART treatment failure.

In this study, it is revealed that being orphan (P=0.049) and residency (P=0.032) was found to be associated with treatment failure. Children who born from HIV positive families with families alive was preventive for treatment failure (AOR=0.51, 95% CI: 0.24-4.77) and moreover, disclosure of HIV status was found to be preventive (AOR=0.61, 95% CI: 0.35-3.29).

## Conclusion

The overall first line ART treatment failure was 18.2% (41patients) in which the most common type is both clinical and immunological (8.9%) followed by immunological failure (6.2%), clinical failure alone accounts for 3.1%. Out of all children who have evidence for first line ART treatment failure, only 14 (34%) patients were detected and started on second line regimens. But about 37(66%) patients were not detected even though they have evidence of treatment failure during follow up. So close monitoring should be done to detect treatment failure early and shift to second line treatment. ARV prophylaxis for PMTCT, advanced clinical stages (3 and 4), base line CD4 less than 200 cells/µl, tuberculosis co-infection, substitution of original regimen once or more for any reason, poor adherence during follow up and duration of ART above 60 months were independent factors that increase the risk of first line ART treatment failure. So this group of patients should be strictly evaluated for treatment failure at every visit since these factors increase the risk of first line ART treatment failure. Service providers should strengthen adherence counseling at every visit since it is a risk factor for treatment failure. Since there are many patients with first line ART treatment failure, second line drugs for children should be available. The trend of shifting to second line treatment and the mean delay from detection of treatment failure and start of second line treatment was not determined in this study and can be studied in subsequent studies.

## Data Availability

The data is available at the Ethiopian Public Health institute Server.

## Declarations

### Ethical considerations

The proposal was reviewed and approved by the scientific and ethical review committee of the EPHI.

### Consent for Publication

There is consent for publication of the scientific findings as part of the consent during data collection

### Availability of data and materials

Data sharing is not applicable to this article as no datasets were generated or analyzed during the current study.

## Acknowledgment

We would like to thank for the EPHI management for the unreserved logistics support. We would also like to acknowledge the Global Fund for financialy supporting this study.

## Abbreviations

AIDS: Acquired immunodeficiency syndrome
ART: Anti-retroviral therapy
DHS: Demographic and Health Surveys
EPHI: Ethiopian Public Health Institute
HFs: Health Facilities
HIV: Human immunodeficiency virus
SERO: Scientific and Ethics Review Office
UNAIDS: Joint United Nations Programme on HIV/AIDS
UNFPA: United Nations Population Fund
VF: Virologic failure
VL: Viral Load
VLS: Viral Load Suppression
WHO: World Health Organization

## References

[1] UNAIDS Report on the global AIDS epidemic | 2012. 2012.

[2] P. Declaration and E. Aids, “Monitoring 2019,” 2019.

[3] T. H. Resources, “Guidelines for the Use of Antiretroviral Agents in Pediatric HIV Infection,” pp. 1–219, 2010.

[4] E. To and F. Hiv, HIV Case Management in Ethiopia: A Pragmatic Approach to Maximizing Adherence to Long-Term Treatment and Retention in Chronic Illness Care..

[5] F. Democratic, “National Guidelines for Comprehensive HIV Prevention, Care and Treatment, 2014,” 2014.

[6] R. September, “National Guidelines for HIV / AIDS and Nutrition,” no. September, 2008.

[7] A. Ababa, “HIV Related Estimates and Projections for Ethiopia March 2018,” no. March, 2018.

[8] A. H. Sohn, “HAART for children with treatment failure,” vol. 3, pp. 485–499, 2009.

[9] J. M. Bernheimer et al., “Paediatric HIV treatment failure : a silent epidemic,” pp. 1–3, 2015.

[10] S. D. Emmett et al., “Predicting Virologic Failure Among HIV-1-Infected Children Receiving Antiretroviral Therapy in Tanzania : a Cross-Sectional Study,” vol. 54, no. 4, pp. 368–375, 2010.

[11] M. R. Kamya et al., “Predictors of Long-Term Viral Failure Among Ugandan Children and Adults Treated With Antiretroviral Therapy,” vol. 46, no. 2, pp. 187–193, 2007.

[12] T. Bacha, B. Tilahun, and A. Worku, “Predictors of treatment failure and time to detection and switching in HIV-infected Ethiopian children receiving first line anti-retroviral therapy,” BMC Infect. Dis., vol. 12, no. 1, p. 1, 2012.

[13] P. Costenaro et al., “Predictors of Treatment Failure in HIV-Positive Children Receiving Combination Antiretroviral Therapy : Cohort Data From Mozambique and Uganda,” vol. 4, no. 1, pp. 39–48, 2015.

[14] T. Rogo, A. K. Delong, P. Chan, and R. Kantor, “Antiretroviral Treatment Failure, Drug Resistance, and Subtype Diversity in the Only Pediatric HIV Clinic in Rhode Island,” vol. 60, 2015.

[15] R. Ginwalla, E. Chama, R. Thomas, M. Mwiya, and C. Kankasa, “Prevalence of Clinical, Immunological and Virological Failure among Children on Haart at the University Teaching Hospital, Lusaka, Zambia,” vol. 39, no. 3, pp. 1–5, 2012.

[16] A. Zeleke, “Prevalence of antiretroviral treatment failure and associated factors in HIV infected children on antiretroviral therapy at Gondar University Hospital, retrospective cohort study,” vol. 8, no. November, pp. 125–132, 2016.

[17] A. Mujugira, C. Celum, J. W. Tappero, A. Ronald, N. Mugo, and J. M. Baeten, “Younger Age Predicts Failure to Achieve Viral Suppression and Virologic Rebound Among HIV-1-Infected Persons,” vol. 32, no. 2, pp. 148–154, 2016.

[18] F. Renaud-théry, B. Nguimfack, M. Vitoria, E. Lee, P. Graaff, and J. Perriëns, “Use of antiretroviral therapy in resource-limited countries in 2006 : distribution and uptake of first- and second-line regimens,” pp. 1–3, 2019.

[19] A. J. Prendergast et al., “Treatment of Young Children with HIV Infection : Using Evidence to Inform Policymakers,” vol. 9, no. 7, pp. 5–10, 2012.

[20] “Table 7. 15 WHO definitions of clinical, immunological and virological failure for the decision to switch ART regimens,” p. 15.

[21] H. I. V. D. Resistance, “Hiv drug resistance,” no. July, 2014.

[22] “World Health Organization Protocol for Cross Sectional Surveillance of Acquired HIV Drug Resistance in Populations Failing First-line Antiretroviral Therapy,” pp. 1–51.

[23] A. Y. Getaneh, A. G. Egziabhier, K. Zealiyas, and R. Tilahun, “Treatment Failure among People living with HIV taking Antiretroviral Therapy in Ethiopia,” vol. 576, 2019.

[24] M. Jonathan et al., “Paediatric HIV Treatment Failure : A Silent Epidemic Authors Paediatric HIV treatment failure : a silent epidemic,” 2019.

[25] R. Sebunya, V. Musiime, S. B. Kitaka, and G. Ndeezi, “Incidence and risk factors for first line anti retroviral treatment failure among Ugandan children attending an urban HIV clinic,” pp. 1–10, 2013.

[26] A. O. Ebonyi et al., “Risk Factors for First-line Antiretroviral Treatment Failure in HIV-1 Infected Children Attending Jos University Teaching Hospital, Jos, North Central Nigeria,” vol. 4, no. 15, pp. 2983–2994, 2014.

[27] O. A. L. Article, “IMMU OLOGIC A D CLI ICAL OUTCOMES OF CHILDRE O HAART : A RETROSPECTIVE COHORT A ALYSIS AT JIMMA U IVERSITY SPECIALIZED HOSPITAL,” no. 6, pp. 75–82, 2006.

[28] O. Article, “Immunological failure despite virological suppression in HIV seropositive individuals on antiretroviral therapy,” vol. 32, no. 2, pp. 94–99, 2011.

[29] J. Kadima et al., “Adoption of routine virologic testing and predictors of virologic failure among HIV-infected children on antiretroviral treatment in western Kenya,” no. Ci, pp. 1–15, 2018.

[30] “Prevalence and risk factors of virological failure among children on antiretroviral therapy,” vol. 2, no. Suppl 2, p. 2017, 2017.

